# Increased persuadability and credulity in people with corpus callosum dysgenesis

**DOI:** 10.1101/2021.12.28.21268413

**Authors:** JM Barnby, R. Dean, H Burgess, J Kim, A.K. Teunisse, L Mackenzie, G Robinson, P Dayan, LJ. Richards

## Abstract

Corpus callosum dysgenesis is one of the most common congenital neurological malformations. Despite being a clear and identifiable structural alteration of the brain’s white matter connectivity, the impact of corpus callosum dysgenesis on cognition and behaviour has remained unclear. Here we build upon past clinical observations in the literature to define the clinical phenotype of corpus callosum dysgenesis better using unadjusted and adjusted group differences compared with a neurotypical sample on a range of social and cognitive measures that have been previously reported to be impacted by a corpus callosum dysgenesis diagnosis. Those with a diagnosis of corpus callosum dysgenesis (*n* = 22) demonstrated significantly higher persuadability, credulity, and insensitivity to social trickery than neurotypical (*n* = 86) participants, after controlling for age, sex, education, autistic-like traits, social intelligence, and general cognition. To explore this further, machine learning, utilising a set neurotypical sample for training the normative covariance structure of our psychometric variables, was used to test whether these dimensions possessed the capability to discriminate between a test-set of neurotypical and corpus callosum dysgenesis participants. We found that participants with a diagnosis of corpus callosum dysgenesis were best classed within dimension space along the same axis as persuadability, credulity, and insensitivity to social trickery after controlling for age and sex, with Leave-One-Out-Cross-Validation across 250 training-set permutations providing a mean accuracy of 71.7%. These results have wide-reaching implications for a) the characterisation of corpus callosum dysgenesis, and b) the role of the corpus callosum in social inference.

## Introduction

Corpus callosum dysgenesis (CCD) is the collective term for congenital abnormalities of the corpus callosum. They include complete (agenesis) or partial absence, as well as a thick or thin (hypoplastic) corpus callosum.^1^ The prevalence of CCD is estimated at 1 to 7 in 4000 live births, making CCD one of the most common congenital neurological malformations^2,3^. While CCD can be detected via ultrasound or neuroradiological imaging from as early as 20 weeks’ gestation^4^, there exist few behavioural phenotypes to qualify this diagnosis.

Individuals with CCD range in clinical presentation from being minimally impacted in cases of isolated CCD to severe cognitive impairment^5^, usually associated with syndromic forms of CCD^1,6,7^. Observational studies have noted difficulties in understanding second order meaning in language, such as proverbs^8,9^, and deficits in executive function, such as difficulties in tap inhibition, cognitive flexibility, decision-making, problem-solving^10,11,12^. One particular class of difficulties reported within CCD are in the domain of social cognition. This includes social insight, social logic, and self-perception^11,13,14^ as well as deficits in inference relating to first- and second order beliefs^14^ and in the interpretation of social intentions^6^. Indeed, difficulties in relationships and navigating the social world have been identified as a critical source of distress in those with a diagnosis of CCD^16^.

Phenotypic features of CCD overlap substantially with those of autism spectrum disorders (ASD)^17^ – a diagnosis with a poorly defined structural neural correlate. This overlap includes over-adherence to social norms^18^, the use of repetitive language, and communication difficulties^19^. Children with CCD exhibit higher incidence rates of scoring above the clinical cut off using the Autism Quotient^20,21^ than the neurotypical (NT) population, with an estimated incidence rate of 44.6% (21/47)^21^ as rated by their parents, implying substantial comorbidity of CCD.

The social difficulties reported in CCD raises the question as to which variations in social functioning are closest to being a hallmark of CCD. Here, we use the covariance amongst psychological characteristics that have previously been noted to be impaired in CCD and autistic individuals to determine whether deficits in social functioning in CCD correlate with the prevalence of autistic-like traits, and whether particular social or cognitive traits in CCD may be statistically dissociable from neurotypical (NT) distributions. This includes social traits such as persuadability and social intelligence, as well as general cognitive traits such as abstract non-verbal reasoning. This will provide additional instruments for understanding the role of defined anatomical deficits in CDD, alongside underpinnings of performance on refined cognitive tasks.

## Materials and Methods

### Participants

Data was collected from two distinct participant cohorts – an NT group, acting as a control, and a set of participants with a CCD diagnosis, as confirmed via magnetic resonance imaging (Figure 1).

**Figure 1.**
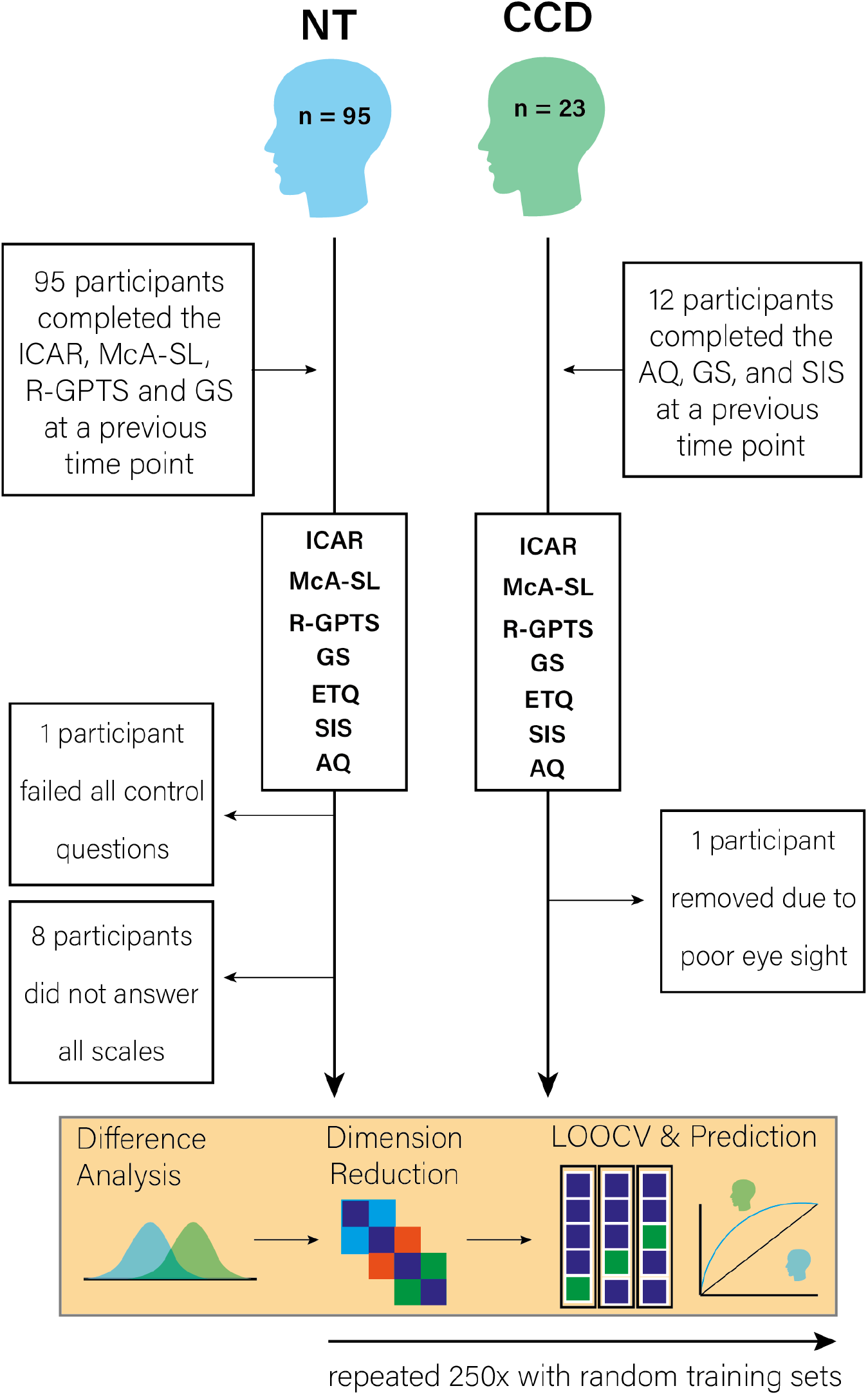
Study design. NT = Neurotypical; CCD = participants with a diagnosis of Corpus Callosum Dysgenesis; ICAR = Matrix Reasoning - International Cognitive Ability Resource; R-GPTS = Revised Green Paranoid Thoughts Scale; McA-SL = MacArthur Social Ladder; GS = Gullibility Scale; AQ = Autism Quotient; ETQ = Epistemic Trust Questionnaire; SIS = Social Intelligence Scale; LOOCV = Leave-One-Out-Cross-Validation.

The CCD cohort consisted of 23 unique participants. These participants were recruited from the Australian Corpus Callosum Dysgenesis Database (a database of individuals with disorders of the corpus callosum and immediate family members, created by the Brain Development and Disorders Laboratory at the University of Queensland) and through partnerships with the family support organisation Australian Disorders of the Corpus Callosum (AusDoCC; https://www.ausdocc.org.au). Data from the CCD group was collected between June and November 2021. Twelve of these participants had also completed the Autism Quotient, Social Intelligence Scale, and Gullibility Scale (see *Measures*) at a previous time point, collected between May and August 2020; this permitted a test-retest reliability analysis to be conducted on measures completed at both time periods by these CCD participants. Those from the CCD cohort who had participated in the 2020 survey were compensated with retail gift vouchers (valued at $15 AUD), with all CCD participants receiving retail gift vouchers (valued at $45 AUD) for their participation in the 2021 survey.

The NT cohort were recruited through the online research platform Prolific Academic. All respondents were between the ages of 18 and 60, had a minimum approval rate of 90% on Prolific Academic and reported no psychiatric or neurological diagnoses, that they resided in Australia, and that they spoke fluent English. During the month of April 2021, 110 NT participants took part in the first round of data collection. Of these 110 participants, 95 (86.3%) completed a second round of data collection during August to September of that same year. All NT participants were paid, through Prolific Academic, £3 for their participation at time point 1 and £5.13 for their participation at time point 2.

One NT participant failed over two checks included in the survey to ensure attentiveness (out of ten), while eight NT participants did not answer all required questions in the survey. These participants were consequently excluded from all analyses, leaving a final sample size of 86. All CCD participants passed all attention checks (out of ten) included in the survey. One CCD participant reported visual difficulties whilst completing the survey, and consequently their data was excluded from all analyses. This resulted in a final sample size of 22 CCD participants, with 12 CCD participants who had repeated measures data available for the test-retest analysis of the Gullibility Scale, and 10 CCD participants who had repeated measures data available for the test-retest analysis of the Social Intelligence Scale and Autism Quotient.

### Measures

#### Experimental Measures

##### Gullibility

Participant gullibility was evaluated using the 12-item Gullibility Scale (GS)^23^. This is comprised of two subscales, each consisting of six questions. The first subscale, Persuadability, measures how readily a person thinks they can be fooled (e.g., ‘My friends think I’m easily fooled; My family thinks I’m a good target for scammers’). The second subscale, Insensitivity, measures the degree to which a participant believes they would be unable to detect social cues as to when they are being manipulated (e.g., ‘I’m pretty poor at working out if someone is tricking me’). Each item is measured on a 7-point scale, with participants rating how strongly they believe each statement from 1 (strongly disagree) to 7 (strongly agree). High scores are taken to be indicative that the participant is likely to be considered socially gullible. This measure was included in this study due to a preliminary analysis of previously collected unpublished pilot data (data not included) and anecdotal reports that those with CCD were more susceptible to social manipulation than their neurotypical contemporaries. Due to the high correlation between these subscales, persuadability and Insensitivity were also combined to create a single ‘Gullibility’ score, in order to facilitate later machine learning analyses (see Dimension Reduction & Prediction).

##### Epistemic Trust

To assess whether those who were more readily persuadable were also generally more trusting and credulous, we administered the Epistemic Trust, Mistrust and Credulity Questionnaire (ETQ)^24^. The ETQ includes three subscales: trust (e.g., ‘I usually ask people for advice when I have a personal problem’), mistrust (e.g., ‘I’d prefer to find things out for myself on the internet rather than asking people for information’), and credulity (e.g., ‘I am often considered naïve because I’ll believe almost anything’). Trust and mistrust have different statistical associations with scales of abuse and neglect,^23^ and therefore have been kept them as separate subscales. Each item is rated on a scale of one (strongly disagree) to seven (strongly agree). Scores on the Trust, Mistrust, and Credulity subscale range from 6-42, with higher scores indicating higher Trust, Mistrust, and Credulity, respectively.

##### Paranoia

Since feelings of paranoia may affect the ability of an individual to intuit the thoughts and intentions of others accurately^25^ the Revised Green Paranoid Thoughts Scale^26^ was also included (R-GPTS). This questionnaire surveys the participant regarding their beliefs about the actions and motivations of others they have interacted with over the past month. This includes two, nine item subscales, one assessing social reference (‘Reference’; e.g., ‘I spent time thinking about friends gossiping about me’) and a second assessing persecutory ideation (‘Persecution’; e.g., ‘People wanted me to feel threatened, so they stared at me’). Each item was scored from 0 (not at all) to four (totally). Previous use of the scale in clinical and non-clinical populations determined cut-off scores: average (Reference: 0–9; Persecution: 0–4), elevated (Reference: 10–15; Persecution: 5–10), moderately severe (Reference: 6–20; Persecution: 11–17), severe (Reference: 21–24; Persecution: 18–17), and very severe (Reference: 25+; Persecution: 28+). Persecutory ideation and social reference subscale scores were combined into a single ‘Paranoia’ score to reduce the effect of collinearity in later machine learning analyses (see Dimension Reduction & Prediction).

##### Relative social standing

The MacArthur Scale of Subjective Social Status (McA-SL; hereafter known as the MacArthur Social Ladder)^27,28^ was used to assess how low or high in the social hierarchy participants view themselves relative to others. It consists of two sub-scales: one for their social ranking within their local community and another for their social ranking nationally. Each scale runs from 0 to 100, with participants being told that 0 means believing that they are ranked at the very bottom of their community/nationally, and 100 means believing that they are ranked at the very top of their community/nationally. The two MacArthur social ladder scales were averaged to generate a mean score for later machine learning analyses, due to their high correlation and identical scale resolution (see Dimension Reduction & Prediction).

#### Control Measures

##### Autism

The prevalence of autistic-like traits in the participant cohorts was assessed using the Autism-Spectrum Quotient (AQ)^20^. The AQ is questionnaire consisting of 50 questions, rated on a 4- point scale, ranging from ‘definitely disagree’ to ‘definitely agree’. The unadjusted and adjusted data analyses to examine differences between group, however, were based on the short version of the AQ (sAQ)^29^, which consists of 28 questions selected from the larger AQ; this was done because this sub-scoring method has been shown to be more reliable and internally consistent across clinical and non-clinical samples.^28^ Possible scores on the sAQ range from 28 to 112, with higher scores indicating that participant has more autistic-like traits. A score of 72 is generally considered a useful cut-off for determining if individuals have a significant level of autistic-like traits,^20^ although this does not in itself fulfil the requirements for a diagnosis of autism.

##### Social intelligence

Social intelligence was assessed using a 21-item questionnaire using the Tromsø Social Intelligence Scale (SIS)^30^ which was translated to English by Grieve and Mahar.^31^ This scale measures three aspects of social intelligence: social information processing (predictions of others’ behaviour), social skills (the ability to interact with others), and social awareness (the detection of social cues). Responses were rated on a 7-point scale from 1 (describes me poorly) to 7 (describes me well). Higher scores indicated higher social intelligence, with scores ranging from 21 – 147.

##### Abstract Non-Verbal Reasoning

We assessed abstract non-verbal reasoning using the progressive matrices task within the Interactive Cognitive Ability Resource (ICAR)^32^. Participants are required to solve ten puzzles in addition to one practice puzzle. In each puzzle, participants are presented with nine shape configurations that follow a specific rule and asked to select a tenth configuration that fitted the rule from six possible answers. The measure was included to control for any general cognitive variance that might otherwise explain differences between and within NT and CCD groups. Scores on this test range from 1-10, with higher scores indicating better abstract non-verbal reasoning.

##### Internal and Test-Retest Reliability of Measures

The internal consistency of each psychometric measure for each group (and at each time point, if applicable) was assessed using Cronbach’s alpha. All measures scored moderate to high internal consistency (α = 0.53 – 0.92; see Table S2 for a full list of each scales value, and previously reported values of each scale). The temporal stability of paranoia, abstract reasoning, subjective social status, and gullibility over time was also assessed across the NT participants. Scales with repeated measures data available calculated from NT had moderate to strong Intraclass Correlation Coefficients (ICC [3,1] = 0.57-0.77)^33^ (Figure S1). ICC analyses of the GS, sAQ, and SIS were conducted using the twelve members of the CCD group who had psychometric scores available at two different time points: persuadability scored excellent reliability (ICC [3,1] = 0.85, p<0.001) and insensitivity scored excellent reliability (ICC [3,1] = 0.91, p<0.001). The sAQ scored excellent reliability (ICC [3,1] = 0.83, p<0.001), as did the R-sAQ (ICC [3,1] = 0.73, p=0.005), and the SIS (ICC [3,1] = 0.79, p=0.002). Given the strong inter-rater reliabilities within the measures that were repeated, all data analyses were conducted on the most recent answers provided by the participant on measures on which they were tested more than once.

### Procedure

All procedures relating to this study were approved by the University of Queensland human research ethics committee (approval ref: 2014/HE000535), and the design of the study itself was created with input from sitting members of the Executive Committee of AusDoCC. All psychometric questionnaires were electronically administered remotely using the Gorilla Experiment Builder for the behavioural sciences^34^, except for the survey administered to the CCD group in 2020, which was electronically administered in the laboratory. The psychometric measures were presented to each participant in a random order, with the order of the questions within each measure similarly randomised. Numeric values and visual indicators were not rendered on a scale until the participant provided an input (via mouse click) to the computer, after which these indicators would become visible.

### Analysis

All analyses were conducted in *R* (4.0.0)^35^ on a Mac OS (Big Sur, 11.5.2; 2.6 GHz 6-core Intel i7).

#### Difference analysis

One-sample t-tests were conducted using the ‘t.test’ base function in R to assess both CCD and NT populations against previously reported means on each scale when raw data from the authors of each original validation paper were not available. Kruskal-Wallis tests were conducted to assess unadjusted group differences for each scale. The ‘psych’^36^ package (2.0.9) was used for descriptive analytics, t-tests, and Kruskal-Wallis analyses.

Given the relatively small CCD sample, and wanting also to control for additional variables such as age and sex in our regression models, we further tested the stability of group differences further using Bayesian regression models which provide uncertainty estimates around each estimated coefficient. Bayesian regression models were fit using the ‘brms’^37^ package, which utilises the ‘Stan’ programming framework for hierarchical Bayesian estimates, and employed a Markov-Chain Monte Carlo (MCMC) method based on a No U-Turn Sampler (NUTS) to draw from the posterior distribution. Each model used four MCMC chains, and each chain ran 2000 iterations (1000 warmup). All models were checked for adequate convergence 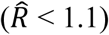 and posterior predictive checks were conducted to ensure the model was able to simulate the true observations adequately.

#### Dimension reduction and prediction

We sought to test whether the psychological phenotype of CCD may be explained by exaggerations to natural psychological variation with NT populations. We included nine variables in this analysis: the ICAR, revised Autism Quotient, Social Intelligence, Gullibility, Trust, Mistrust, Credulity, Paranoia, and the McA-SL. As the sAQ contains several items that overlapped with the Social Intelligence Scale, the decision was made to remove these items and create a Revised sAQ sub-scale to avoid collinearity in the analysis (R-sAQ; see Supplementary Materials; Table S1). We then regressed out the influence of age and sex on each variable across both NT and CCD data sets using linear models. The resulting residuals of these regressions for each variable were then used for the dimension reduction and prediction.

We first conducted unsupervised exploratory dimension reduction on a random training set of NT data (*n* = 60, ~70%) using the ‘factoextra’^38^ package. This uses the principal components of the data extracted using the ‘prcomp’ function to assess the latent structure that may explain variance between multiple measures. We retained dimensions explaining more than 10% of the variance. We used these top dimensions obtained from the training data to predict whether a participant was a member of either the CCD or NT group using the left-out test participants in the NT sample (*n* = 26) and all participants in the CCD (*n* = 22) group. Predictions were made using Leave-One-Out-Cross-Validation (LOOCV) using a Bayesian General Linear Model classifier. In addition, we extracted a confusion matrix from the LOOCV.

To ensure our dimension reduction and prediction procedure was not biased to a particular training set, we repeated the above procedure 250 times using random NT training and test data on each repetition (Training *n* = 60; ~70%; Test *n* = 26; ~30%). We then present the average variance explained and loading of variables on each of the top dimensions from the principal component analysis, the average accuracy of the LOOCV models, and averaged confusion matrices across all 250 repetitions.

## Data availability

All anonymised data and analysis code are available for free from https://github.com/Brain-Development-and-Disorders-Lab.

## Results

The NT participants were relatively young, mostly male, and mostly educated at the undergraduate level (see Table S1). The CCD participants were significantly older than NT participants (CCD mean[sd] age = 47 [17.26]; NT mean[sd] age = 29.4 [7.60]; t(25.01) = 5.14, 95%CI: 9.90, 23.13; ***P* <0.001**), were mostly female (CCD = 59.1% female; NT = 40.2% female) and were mostly educated at the high school level (see Table S1).

### Comparison with previously reported data

The NT participants had ICAR scores that were above average compared to previously studied general population samples when assessing by average item score^32^ (t(85) = 5.45, *P*<0.001; average item score = 0.52), whereas members of the CCD cohort were not significantly different from previously reported means in the general population (t(22) = - 0.56, *P*=0.57).

Both NT and CCD participants were on the upper end of ‘average’ to the lower end of ‘elevated’ regarding both their social reference and persecutory ideation scores when compared with validated cut-offs derived from non-clinical and clinical populations.^26^ The means of both groups were significantly below the clinical cut offs for the persecution subscale (NT = (t(85) = -11.98, *P* <0.001); CCD = (t(22) = -3.28, *P* =0.004)).

Considering gullibility, the NT participants were significantly lower in persuadability (t(85) = -3.84, *P* <0.001) but not higher in insensitivity (t(85) = 0.03, *P* =0.98), while those with CCD were elevated in both persuadability (t(22) = 2.81, *P* =0.010) and insensitivity (t(22) = 3.23, *P* =0.004) relative to general population data from two prior studies.^23^

The original study that validated the Epistemic Trust measure^24^ used the average item score for each participant and each subscale (from 1-7) for assessment; the authors have kindly made their data available for full group comparisons online^24^. Using the same assessment of participant scores for each subscale, our NT group were not significantly different from their sample in trust (t(93.97) = -1.18, *P* = 0.23), although were slightly higher than previous data in mistrust (t(96.68) = 2.19, *P* = 0.03) and slightly lower in credulity than previous data (t(100.33) = -3.08, *P* = 0.003). By contrast, our CCD participants were significantly higher in trust than the original study (t(21.78) = 2.98, *P* = 0.007), were not significantly lower in mistrust (t(21.60) = 0.16, *P* = 0.87), and were significantly higher in credulity (t(21.43) = 2.64, *P* = 0.015).

By assessing the mean of the prior data reported from the sAQ^29^ from neurotypical participants (mean = 56.74), it was determined that our NT participants scored significantly above the prior mean (t(85) = 8.12, *P* < 0.001) as did the CCD participants (t(21) = 8.50, *P* < 0.001). Additionally, by comparing against the mean of previously reported participants with a diagnosis of Asperger’s syndrome (85.63), it was found that our NT participant scored significantly below the prior mean (t(85) = -22.52, *P* < 0.001), as did our CCD participants (t(21) = -6.26, *P* < 0.001).

Finally, comparing against the previously reported means of the SIS^30^ (mean = 102.62), we find that our NT participants were significantly below the prior mean (t(85) = -3.13, *P* = 0.002), as were our CCD participants (t(21) = -5.05, *P* < 0.001).

### Group differences

Simple unadjusted Wilcoxon-rank sum tests of difference between NT and CCD participants (Figure 2) showed that those with CCD were on average lower in abstract non-verbal intelligence (ICAR; χ^2^(1) = 10.06, *P* = 0.002). Those with CCD were more persuadable (χ^2^(1) = 14.52, *P* < 0.001), and more insensitive to social cues of trickery (χ^2^(1) = 8.76, *P* = 0.003), more trusting (χ^2^(1) = 6.39, *P* = 0.011), and scored higher on credulity (χ^2^(1) = 13.41, *P*< 0.001). CCD participants also scored higher in autistic traits (χ^2^(1) = 12.36, *P* < 0.001) and lower on social intelligence (χ^2^(1) = -7.65, *P* = 0.006) compared to NT participants. Those with CCD did not perceive themselves to be any higher or lower than NTs on either of the MacArthur Social Ladders (National Ladder: χ^2^(1) = 0.11, *P* = 0.75; Community Ladder: χ^2^(1) = 0.25, *P* = 0.62), nor were any less mistrusting than NT participants (χ^2^(1) = 0.92, *P* = 0.34). There was no difference between groups for persecutory ideation (χ^2^(1) = 0.30, 0.58) or social reference (χ^2^(1) = 0.69, *P* = 0.41).

**Figure 2.**
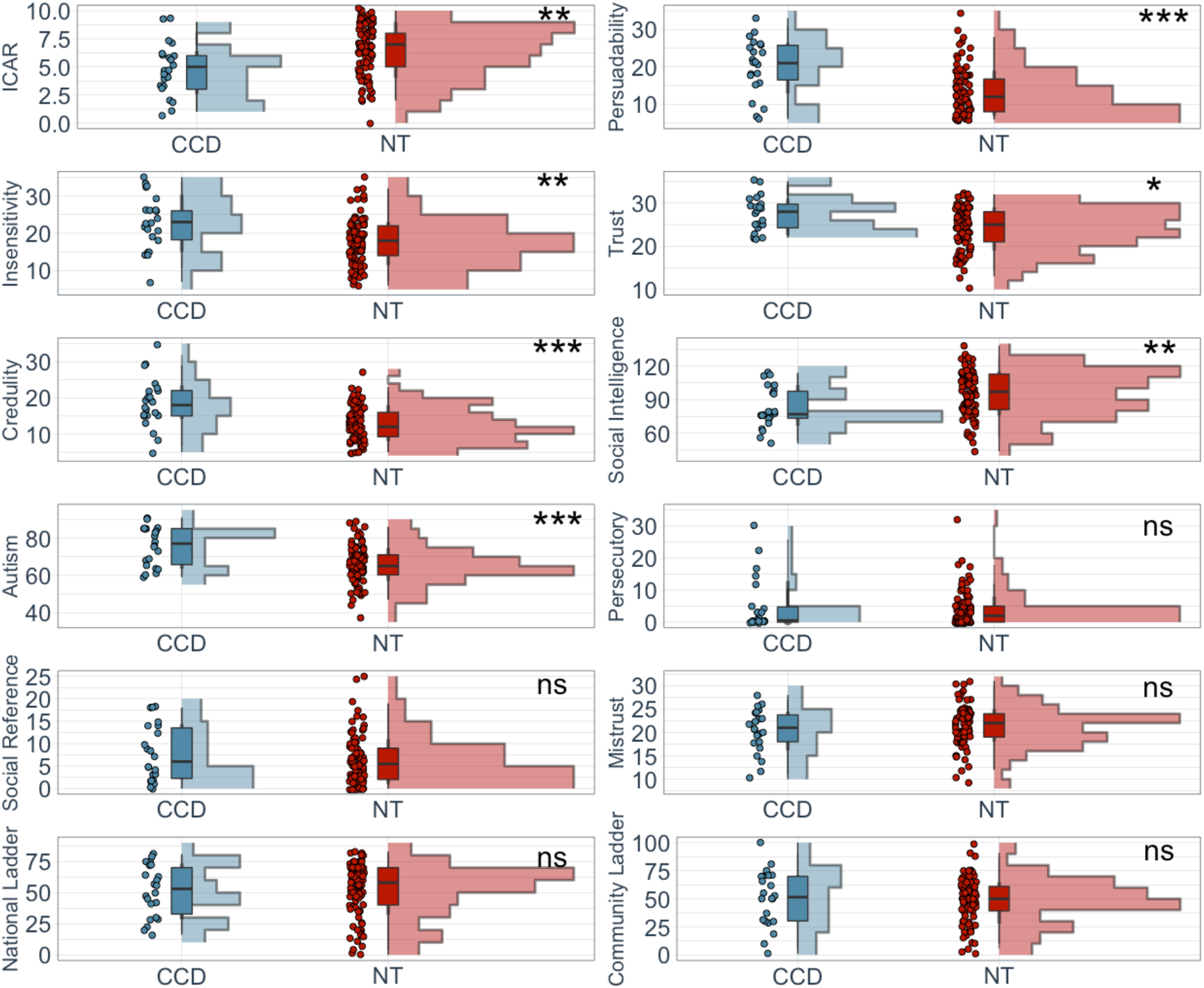
Unadjusted Kruskal-Wallis tests between CCD and NT participants for each psychometric measure. Those with CCD were on average lower in general cognitive ability and social intelligence, and higher on persuadability, insensitivity to social trickery, trust, credulity, and autistic-like traits compared to neurotypical participants. Adjusting for age, sex, and education however removed the difference in general cognition, and adjusting for age, sex, education, and general cognition removed the difference in social intelligence. Points on each graph represent individual scores for each measure within each group. Boxplots represent the median and quartile range. Histograms represent the density of points for each group at each score on the scale as a function of sample size. ns = not significant; * = p < 0.05; ** = p < 0.01; *** = p < 0.001.

Bayesian general linear models were employed to examine these relationships further (and quantify estimate noise) and account for group-based differences in age, sex, and education. The results of this analysis indicate that the CCD participants were no longer lower in abstract non-verbal intelligence (median posterior estimate (*m*): -1.30, 95% Credible Interval (95%CI): -2.71, 0.10).

Delving further into individual items on the ICAR, mixed Bayesian regression modelling of reaction time between CCD and NT groups, with ‘(Question Number | ID)’ as a random variable, demonstrated that NT participants were significantly quicker at answering questions (*m*: -15571.22, 95%CI: -30066.21, -1446.41; see Figure S3) regardless of whether their answers were correct or incorrect. Correct answers were not slower overall (*m*: 2070.80, 95%CI: -7318.45, 11901.53) and there was no interaction between group and whether an answer was answered correctly or incorrectly on reaction time (*m*: -5782.85, 95%CI: - 16794.47, 5604.07). Likewise, by regressing normative item difficulty (previously defined for the ICAR progressive matrices^39^) and group (with ID as a random variable) against whether participants got an item correct or not, we observed that increasing item difficulty led to fewer correct answers regardless of group identification (*m*: -1.76, 95%CI: -2.41, - 1.12), and that there was an interaction between group and difficulty, such that those with CCD provided significantly fewer correct answers as difficulty increased compared to controls (*m*: -0.78, 95%CI: -1.49, -0.09). As a result of this finding, the participant’s average ICAR score was included in all future models, alongside age, sex, and educational attainment, to assess whether psychometric effects may be explained by lower abstract non-verbal intelligence.

All significant effects from the simple group comparisons remained when controlling for the ICAR, age, sex, and education: CCD participants were more persuadable (*m*: 9.76, 95%CI: 5.67, 13.66), more insensitive to noticing social trickery (*m*: 7.39, 95%CI: 3.40, 11.47), more trusting (*m*: 4.52, 95%CI: 1.61, 7.46) and had higher credulity (*m*: 7.68, 95%CI: 4.61, 10.80). CCD participants also scored higher in autistic traits (*m*: 8.53, 95%CI: 2.15, 14.92), although were no longer lower on social intelligence (*m*: -9.59, 95%CI: -22.35, 2.86) compared to NT. Those with a diagnosis of CCD were still no different to NT participants in mistrust (*m*: - 1.76, 95%CI: -4.49, 0.88), social reference (*m*: 3.22, 95%CI: -0.15, 6.45), or persecutory ideation (m: 0.11, 95CI: -3.79, 3.66), or on the MacArthur social ladders.

When additionally including autism and social intelligence alongside age, sex, education, and ICAR score in a Bayesian general linear model predicting gullibility subscales, CCD participants still scored higher in persuadability (*m*: 8.43, 95%CI: 4.47, 12.45) and insensitivity (*m*: 6.51, 95%CI: 2.95, 10.03). In other words, differences in age, sex, education, general cognition, autistic traits, and social intelligence accounted for less variability in the linear regression model than having a diagnosis of CCD.

### Dimension reduction and prediction

Spearman correlation matrices for the scales were used during the dimension reduction in each group (R-GPTS, AQ, SIS, McA-SL, ETQ, GQ), in addition to calculating whether associations were significantly different between groups (Figure S3). There was only one significant difference in the correlation matrices between groups; a significant positive association between Credulity and Mistrust (that existed for both the NT group, *r*_*spearman*_ = 0.21, *P* = 0.047, and the CCD group, *r*_*spearman*_ = 0.68, *P* = 0.01) which significantly differed from each other following permutation testing (10000 sampled repetitions: *P* = 0.021).

The dimension reduction and prediction modelling were conducted using 250 repetitions, each drawing on randomly sampled training data per repetition (*n* = 60; ~70%). Three principal components were identified, which each explained more than 10% of the variance in the model, with the three components together explaining 62.2% of the variance in total (PC1_%_ = 34.5, 95%CI: 34.2, 34.8; PC2_%_ = 15.6, 95%CI: 15.5, 15.7; PC3_%_ = 12.1, 95%CI: 12.0, 12.2). Upon reviewing the averaged absolute covariance matrices from all 250 training permutations over a threshold of 0.30 (Figure S3A), and the contribution of variables within dimension space (Figure 3D & 3E), absolute loading values above the threshold were generated for gullibility and credulity that loaded jointly on PC1 and PC2 (PC1: Gullibility = 0.410 95%CI: 0.407, 0.413; Credulity = 0.413, 95%CI: 0.409, 0.418; PC2: Gullibility = 0.332, 95%CI: 0.319, 0.346; Credulity = 0.407, 95%CI: 0.396, 0.419) and trust with PC2 and PC3 dimensions (PC2 = 0.594, 95%CI: 0.580, 0.608; PC3 = 0.380, 95%CI: 0.357, 0.404) while all other variables were only above the loading threshold with either PC1, PC2, or PC3 (see Figure 3A).

**Figure 3:**
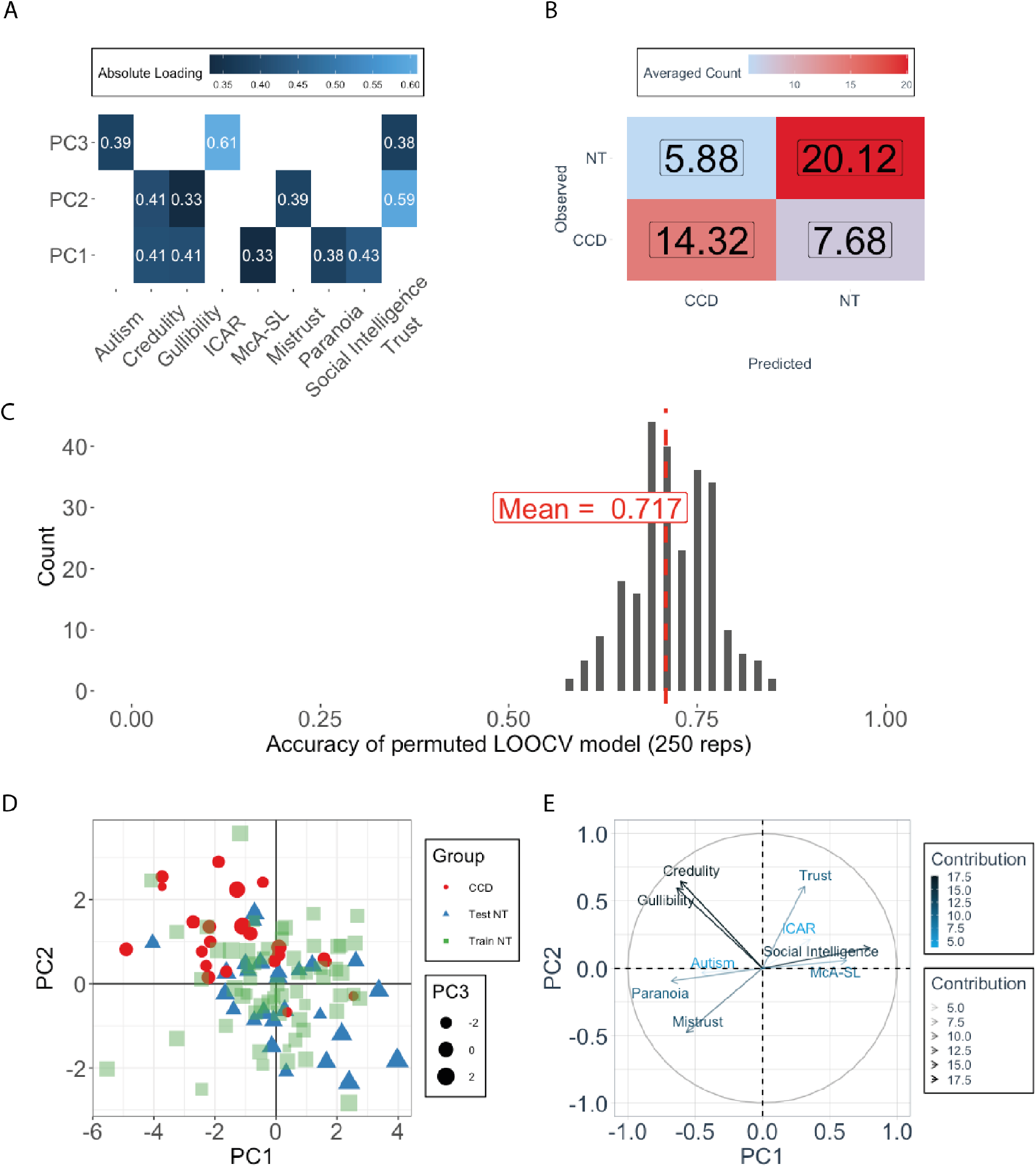
Dimension rotation and LOOCV results across 250 repetitions. (A) Averaged absolute loading between each variable and the top three principal components over all 250 repetitions. All values < 0.30 are filtered out. (B) Averaged permuted confusion matrix (250 repetitions) for each Leave-one-out-cross-validation analysis using a Bayesian GLM classifier. X = non-significant association (alpha = 0.05). (C) Permuted LOOCV model accuracy scores (250 repetitions) using a random 70:30 split of training and test NT data. (D) Projected location in dimension space of test-set NT participants (blue triangles) and CCD participants (red dots) when considered within a random repetition from dimension space built on training set NT participants (green squares). (E) Loading and contribution of each variable to the top two dimensions from the same random repetition as (D).

The top three principal components extracted from dimension reduction were used in a LOOCV to predict group classification (CCD or NT) in the held-out NT data and CCD data in each repetition. An averaged confusion matrix was extracted for predicted and observed classifications across all 250 LOOCV models (Figure 3B), which provided a mean accuracy of 71.7% (95%CI: 71.1, 72.4; Figure 3C).

## Discussion

CCD is easily identified by structural imaging, but the corresponding behavioural phenotype is unclear and highly heterogenous. In this exploratory study we assessed the psychometric profile of those with CCD compared to NT participants. Motivated by prior work on the social difficulties reported in CCD^6,11,13,14,16^ and high comorbidity of ASD in this population^18^, the focus of this work was on identifying any consistent psychometric differences that could not be explained by age, sex, education, poorer general cognitive ability, or autistic traits. It was determined that persuadability, insensitivity to noticing social trickery, credulity, trust, and autistic traits were significantly higher in those with CCD compared to NT populations following adjustment by Bayesian general linear models. To explore this distinction further, a machine learning model built from a training set of NT participants was used to test whether it was possible to classify whether a participant was part of the CCD sample. The result of this analysis was that a normative model built from cognitive and social variables drawing on NT data is reasonably predictive of classifying the CCD phenotype, which is supportive of the theory that there is a consistent psychological phenotype associated with CCD.

Traditionally, previous attempts to define a clear psychological phenotype for CCD has been a difficult challenge – cognitive and behavioural expressions in CCD are highly heterogenous, and previous works have typically used small samples to draw observations.^6^ The covariance structure extracted from the dimension reduction suggests that those diagnosed with CCD are primarily typified by exaggerations in persuadability, credulity, and insensitivity to social trickery. Those with lived experienced of CCD have anecdotally reported their difficulties in understanding the social intentions of others, and it may be that a predisposition to be overly trusting has led some of those with CCD into abusive relationships or finding themselves being the target of antisocial behaviours, such as bullying^16^. More broadly, this highlights the necessity of having CCD recognised as a vulnerable group in healthcare policy so that adequate social and financial support can be arranged. While we do not have sufficient power in this study to form a fully realised, clinically validated model for diagnostic utility, this preliminary model may be useful to examine more nuanced changes to the corpus callosum (e.g., thickness, functional activity) and its relationship with different dimensions of cognition. It will be important to replicate these findings in a new CCD sample, as well as in those who have a clinical diagnosis of ASD without CCD, and individuals who have had their corpus callosum surgically severed as an adult (i.e., callosotomised, or split-brain, individuals given their similar structural changes but different behavioural phenotype)^40^ to assess the specificity of the findings reported here to those with CCD.

The results of this work also demonstrate that persuadability, credulity, and insensitivity to social trickery are statistically dissociable from the other general and social cognitive scales we included in this analysis within the general population. This is consistent with parallel work in field of hypnosis research, regarding the trait of ‘suggestibility’. Phenomenologically consistent with gullibility, especially ‘secondary suggestibility’,^41^ suggestibility has been found to share little to no relationship with canonical personality dimensions defined in Big 5 or OCEAN, such as agreeableness and neuroticism.^42-45^ Indeed, suggestibility has been considered an independent cognitive facet,^45^ and has demonstrated a high degree of stability, persisting over a period of 25 years in the same group of individuals.^47^ Participant responsiveness to direct verbal suggestions (i.e., ‘Direct Verbal Suggestibility’, or DVS^46,48,49^, with and without hypnosis, was outside the scope of this work and so it remains unclear whether exaggerated persuadability and insensitivity to social trickery in our CCD samples will translate into DVS. Rigorously exploring the possibility that CCD relates to both gullibility (second order suggestibility) and DVS in the future works may highlight an important role of the corpus callosum in this separable social trait, and work toward a clearer phenotype of the diagnosis.

Our finding that those with CCD are significantly more likely to score higher on autistic-like traits than the general population is consistent with previous work showing that a large minority of children diagnosed with CCD are also comorbid with ASD.^22^ It has been hypothesised that CCD may be a risk factor in the development of ASD and may explain many of the social difficulties observed in CCD^19^. However, prior work has also found that after parent-rated assessments were included, many of the ASD scores in those with CCD were attenuated below the clinical threshold^19^. As qualitative work has identified the preference for many individuals with CCD to be alone because of negative social experience, such as ostracism, abuse, and social anxiety^16^, it may be that many of the autistic-like qualities that the sAQ aims to measure are confounded by these learnt social responses following social exclusion, rather than CCD sharing many biologically causal links with ASD. Indeed, we find that even after controlling for ASD-like traits and social intelligence, exaggerations in credulity, insensitivity to social trickery, and persuadability remain. Therefore, while phenomenologically CCD and ASD have multiple similarities, their biological, social, and psychological causes may be very different.

Finally, this study corroborates our previous work indicating that CCD impacts abstract non-verbal reasoning^50^. In a Latin Square Task, it was found that increasing task complexity led to more reasoning errors and increased reaction times^50^. Our sample of CCD individuals were no different from our NT sample in terms of their overall ICAR scores when age, sex, and education was controlled for; however, those with CCD were more prone to incorrectly answering more complex items on the ICAR (as defined by Subotic and colleagues)^39^ wrongly when compared to healthy controls, even when considering item difficulty as an independent factor in whether a participant answered an item correctly or not. This supports the hypothesis that those with CCD are more prone to errors when faced with increasing complexity in abstract non-verbal reasoning. However, this hypothesis requires further testing with cognitive tasks that require the navigation of non-verbal environments, such as the assessment of model-free and model-based reasoning using symbolic stimuli^51^.

A limitation of the current study is that individuals with CCD may exhibit reductions in metacognitive efficiency (data not reported here) and therefore gathering data from self-reflective scales alone may not be accurate. In future work, it may prove useful to obtain family-rated and clinician-rated behaviours of the participants to confirm self-reported scores. It should be noted however that the same issue extends to the NT participants. Likewise, it has been noted that collecting data online using psychometric self-report measures can incur spurious results^52^. This was controlled for, in part, by having both the NT and CCD groups complete periodic attention and comprehension checks. However, the utilisation of online testing for those with CCD does offer certain advantages over lab-based observation. For example, those with CCD commonly experience high social anxiety and unease in unfamiliar environments, such as attending a laboratory session for in-person testing. Furthermore, online-testing permits geographically isolated individuals, either by their physical distance from the laboratory or public health restrictions, to participate and gather important cognitive and phenomenological data in a time and cost-efficient manner. It is imperative, however, that appropriate controls (reverse coded items, attention check questions, comprehension questions) are given alongside any surveys and tasks that are administered remotely to those all participants to ensure self-report and online-tested cognitive tasks are reliable. Finally, while the CCD cohort was representative of a broad range of cognitive capabilities, this did not extend to those with severe intellectual disabilities. Therefore, the primary findings of this work may not readily generalise to those who are profoundly impaired.

In summary, the psychometric dimensions of those with CCD were assessed in comparison with NT populations. It was identified that those with CCD demonstrate exaggerated vulnerabilities to social persuasion and are less aware of their possibly being deceived. Machine learning approaches identified that persuadability and insensitivity to social trickery were dissociable elements in the NT population, and the resultant covariance structure was able to accurately predict whether test participants belonged to the CCD or NT group. It is our hope that these results can be used as a foundation on which to develop a more robust model to predict CCD phenomenology, as well as expound upon the contributions the corpus callosum makes to social inference.

## Abbreviations

AQ: Autism Quotient
ASD: Autism Spectrum Disorder
CCD: Corpus Callosum Dysgenesis
ETQ: Epistemic Trust Questionnaire
GS: Gullibility Scale
ICAR: International Cognitive Ability Resource (Progressive Matrices)
ICC: Intraclass Correlation Coefficient
LOOCV: Leave-One-Out-Cross-Validation
McA-SL: MacArthur Social Ladder
NT: Neurotypical
PC: Principal Component
R-GPTS: Revised Green Paranoid Thoughts Scale
SIS: Social Intelligence Scale.

## Acknowledgements

We would like to thank the members of AusDoCC for their co-production in the development of this study, and their unwavering support and commitment to the understanding of CCD. Without their collaboration our work would not exist.

## References

1. Edwards, T. J., Sherr, E. H., Barkovich, A. J., & Richards, L. J. (2014). Clinical, genetic, and imaging findings identify new causes for corpus callosum development syndromes. Brain, 137(6), 1579–1613.

2. Glass, H. C., Shaw, G. M., Ma, C., & Sherr, E. H. (2008). Agenesis of the corpus callosum in California 1983–2003: A population-based study. American Journal of Medical Genetics Part A, 146(19), 2495–2500.

3. Wang, L. W., Huang, C. C., & Yeh, T. F. (2004). Major brain lesions detected on sonographic screening of apparently normal term neonates. Neuroradiology, 46(5), 368–373.

4. Pooh RK. (2009), Neuroscan of congenital brain abnormality, pp 60–139 in Fetal Neurology, Edited by Ritsuko K Pooh and Asim Kurjak, Jaypee Brothers Medical Publishers.

5. Spencer-Smith, M., Knight, J.L., Lacaze, E., Irc5 Consortium, Depienne, C., Lockhart, P.J., Richards, L.J., Heron, D., Leventer, R.J., & Robinson, G.A. 2020. Callosal agenesis and congenital mirror movements: outcomes associated with DCC mutations. Developmental Medicine & Child Neurology, 62(6), pp.758–762.

6. Brown, W. S., & Paul, L. K. (2019). The neuropsychological syndrome of agenesis of the corpus callosum. Journal of the International Neuropsychological Society, 25(3), 324–330.

7. Paul, L. K., Brown, W. S., Adolphs, R., et al. (2007). Agenesis of the corpus callosum: genetic, developmental, and functional aspects of connectivity. Nature Reviews Neuroscience, 8(4), 287–299.

8. Brown, W. S., Paul, L. K., Symington, M., & Dietrich, R. (2005). Comprehension of humor in primary agenesis of the corpus callosum. Neuropsychologia, 43(6), 906–916.

9. Rehmel, J. L., Brown, W. S., & Paul, L. K. (2016). Proverb comprehension in individuals with agenesis of the corpus callosum. Brain and Language, 160, 21–29.

10. Brown, W. S., Anderson, L. B., Symington, M. F., & Paul, L. K. (2012). Decision-making in individuals with agenesis of the corpus callosum: Expectancy-valence in the Iowa Gambling Task. Archives of clinical neuropsychology, 27(5), 532–544.

11. Brown, W. S., & Paul, L. K. (2000). Cognitive and psychosocial deficits in agenesis of the corpus callosum with normal intelligence. Cognitive neuropsychiatry, 5(2), 135–157.

12. Marco, E. J., Harrell, K. M., Brown, W. S., et al. (2012). Processing speed delays contribute to executive function deficits in individuals with agenesis of the corpus callosum. Journal of the International Neuropsychological Society, 18(3), 521–529.

13. Kosky, K. M., Phenis, R., & Kiselica, A. M. (2021). Neuropsychological functioning in dysgenesis of the corpus callosum with colpocephaly. Applied Neuropsychology: Adult, 1-7.

14. Symington, S. H., Paul, L. K., Symington, M. F., Ono, M., & Brown, W. S. (2010). Social cognition in individuals with agenesis of the corpus callosum. Social Neuroscience, 5(3), 296–308.

15. Melogno, S., Pinto, M. A., Scalisi, T. G., Badolato, F., & Parisi, P. (2021). Case Report: Theory of Mind and Figurative Language in a Child with Agenesis of the Corpus Callosum. Frontiers in Psychology, 11:596804.

16. Maxfield, M., Cooper, M. S., Kavanagh, A., Devine, A., & Gill-Atkinson, L. (2021). On The Outside Looking In: A Phenomenological Study of The Lived Experience of Australian Adults with A Disorder of The Corpus Callosum.

17. Demopoulos, C., Arroyo, M. S., Dunn, W., Strominger, Z., Sherr, E. H., & Marco, E. (2015). Individuals with agenesis of the corpus callosum show sensory processing differences as measured by the sensory profile. Neuropsychology, 29(5), 751–758.

18. Brown, W. S., Burnett, K. A., Vaillancourt, A., & Paul, L. K. (2021). Appreciation of social norms in agenesis of the corpus callosum. Archives of Clinical Neuropsychology. 36(7), 1367–1373.

19. Paul, L. K., Corsello, C., Kennedy, D. P., & Adolphs, R. (2014). Agenesis of the corpus callosum and autism: a comprehensive comparison. Brain, 137(6), 1813–1829.

20. Baron-Cohen, S., Wheelwright, S., Skinner, R., Martin, J., & Clubley, E. (2001). The autism-spectrum quotient (AQ): Evidence from Asperger syndrome/high-functioning autism, males and females, scientists, and mathematicians. Journal of Autism and Developmental Disorders, 31(1), 5–17.

21. Baron-Cohen, S., Hoekstra, R. A., Knickmeyer, R., & Wheelwright, S. (2006). The autism-spectrum quotient (AQ)—adolescent version. Journal of Autism and Developmental Disorders, 36(3), 343–350.

22. Lau, Y. C., Hinkley, L. B., Bukshpun, P., et al. (2013). Autism traits in individuals with agenesis of the corpus callosum. Journal of Autism and Developmental Disorders, 43(5), 1106–1118.

23. Teunisse, A. K., Case, T. I., Fitness, J., & Sweller, N. (2020). I Should have known better: Development of a self-report measure of gullibility. Personality and Social Psychology Bulletin, 46(3), 408–423.

24. Campbell, C., Tanzer, M., Saunders, R., et al. (2021). Development and validation of a self-report measure of epistemic trust. PloS one, 16(4), e0250264.

25. Fletcher, P. C., & Frith, C. D. (2009). Perceiving is believing: a Bayesian approach to explaining the positive symptoms of schizophrenia. Nature Reviews Neuroscience, 10(1), 48–58.

26. Freeman, D., Loe, B. S., Kingdon, D., et al. (2021). The revised Green et al., Paranoid Thoughts Scale (R-GPTS): psychometric properties, severity ranges, and clinical cut-offs. Psychological Medicine, 51(2), 244–253.

27. Adler, N. E., Epel, E. S., Castellazzo, G., & Ickovics, J. R. (2000). Relationship of subjective and objective social status with psychological and physiological functioning: Preliminary data in healthy, white women. Health Psychology, 19(6), 586–592.

28. Goodman, E., Adler, N. E., Daniels, S. R., Morrison, J. A., Slap, G. B., & Dolan, L. M. (2003). Impact of objective and subjective social status on obesity in a biracial cohort of adolescents. Obesity research, 11(8), 1018–1026.

29. Hoekstra, R. A., Vinkhuyzen, A. A., Wheelwright, S., et al. (2011). The construction and validation of an abridged version of the autism-spectrum quotient (AQ-Short). Journal of autism and developmental disorders, 41(5), 589–596.

30. Silvera, D., Martinussen, M., & Dahl, T. I. (2001). The Tromsø Social Intelligence Scale, a self-report measure of social intelligence. Scandinavian Journal of Psychology, 42(4), 313–319.

31. Grieve, R., & Mahar, D. (2013). Can social intelligence be measured? Psychometric properties of the Tromsø Social Intelligence Scale–English version. The Irish Journal of Psychology, 34(1), 1–12.

32. Condon, D. M., & Revelle, W. (2014). The international cognitive ability resource: Development and initial validation of a public-domain measure. Intelligence, 43, 52–64.

33. Koo, T. K., & Li, M. Y. (2016). A guideline of selecting and reporting intraclass correlation coefficients for reliability research. Journal of Chiropractic Medicine, 15(2), 155–163.

34. Anwyl-Irvine, A. L., Massonnié, J., Flitton, A., Kirkham, N., & Evershed, J. K. (2020). Gorilla in our midst: An online behavioral experiment builder. Behavior Research Methods, 52(1), 388–407.

35. R Core Team (2020). R: A language and environment for statistical computing.

36. Revelle, R (2020). Procedures for Psychological, Psychometric, and Personality Research. v. 2.0.9

37. Bürkner, P. C. (2017). brms: An R package for Bayesian multilevel models using Stan. Journal of Statistical Software, 80(1), 1–28.

38. Kassambra & Mundt (2020) Extract and Visualize the Results of Multivariate Data Analyses. v. 1.0.7

39. Subotić, S., Baošić, J., Velimir, A., MalenǍić, M., Mihajlović, B., & Lovrić, S. R. (2020). Psychometric validation of the ICAR Matrix Reasoning test. Empirical Studies in Psychology, 59–62.

40. Siffredi, V., Anderson, V., Leventer, R. J., & Spencer-Smith, M. M. (2013). Neuropsychological profile of agenesis of the corpus callosum: a systematic review. Developmental Neuropsychology, 38(1), 36–57.

41. Eysenck, H. J., & Furneaux, W. D. (1945). Primary and secondary suggestibility: an experimental and statistical study. Journal of Experimental Psychology, 35(6), 485.

42. Gudjonsson, G. H. (1983). Suggestibility, intelligence, memory recall and personality: An experimental study. The British Journal of Psychiatry, 142(1), 35–37.

43. Liebman, J. I., McKinley-Pace, M. J., Leonard, A. M., et al. (2002). Cognitive and psychosocial correlates of adults’ eyewitness accuracy and suggestibility. Personality and Individual Differences, 33(1), 49–66.

44. Norris, D. L. (1973). Barber’s task-motivational theory and post-hypnotic amnesia. American Journal of Clinical Hypnosis, 15(3), 181–190.

45. Pires, R., Silva, D. R., & Ferreira, A. S. (2013). Personality styles and suggestibility: A differential approach. Personality and Individual Differences, 55(4), 381–386.

46. Oakley, D. A., Walsh, E., Mehta, M. A., Halligan, P. W., & Deeley, Q. (2021). Direct verbal suggestibility: Measurement and significance. Consciousness and Cognition, 89, 103036.

47. Piccione, C., Hilgard, E. R., & Zimbardo, P. G. (1989). On the degree of stability of measured hypnotizability over a 25-year period. Journal of Personality and Social Psychology, 56(2), 289–295.

48. Oakley, D. A., Walsh, E., Lillelokken, A. M., Halligan, P. W., Mehta, M. A., & Deeley, Q. (2020). United Kingdom norms for the harvard group scale of hypnotic susceptibility, form A. International Journal of Clinical and Experimental Hypnosis, 68(1), 80–104.

49. Oakley, D. A., & Halligan, P. W. (2017). Chasing the rainbow: the non-conscious nature of being. Frontiers in psychology, 8, 1924.

50. Hearne, L. J., Dean, R. J., Robinson, G. A., Richards, L. J., Mattingley, J. B., & Cocchi, L. (2019). Increased cognitive complexity reveals abnormal brain network activity in individuals with corpus callosum dysgenesis. NeuroImage: Clinical, 21, 101595.

51. Keramati, M., Smittenaar, P., Dolan, R. J., & Dayan, P. (2016). Adaptive integration of habits into depth-limited planning defines a habitual-goal–directed spectrum. Proceedings of the National Academy of Sciences, 113(45), 12868–12873.

52. Zorowitz, S., Niv, Y., & Bennett, D. (2021). Inattentive responding can induce spurious associations between task behavior and symptom measures. PsyArXiv. https://doi.org/10.31234/osf.io/rynhk

