## Supplementary Figures and Tables for "Increased persuadability and credulity in people with corpus callosum dysgenesis"

### Supplementary Material

#### Figure S1 Intraclass Correlation Coefficient analysis

Intraclass correlation coefficient and 95% confidence intervals for each subscale within each measure. Koo & Li (2016) define scores between 0.50 and 0.75 as moderate, and between 0.75 and 0.90 as good.

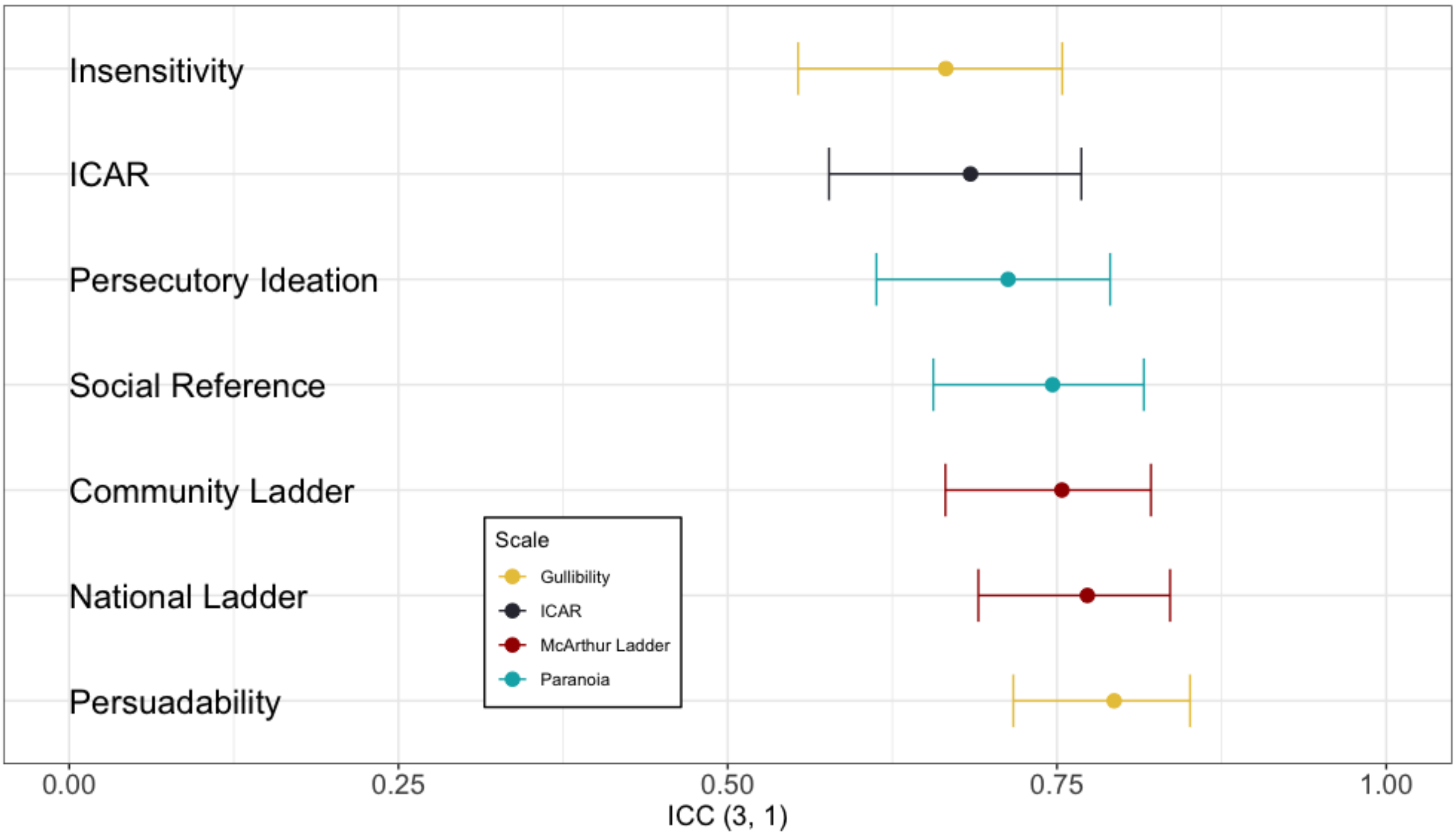

### Figure S2 Psychometric properties of the ICAR for NT and CCD participants

(A) Association between normative question difficulty (Subotic et al., 2020) and correct answers on the ICAR progressive matrices. Points represent the true observed predictions of whether a participant got a question correct (1) or incorrect (0) and the line denotes the logistic function predicting the probability of a correct answer given a range of difficulties from -0.19 – 1.72 in steps of 0.1, modelled on the observed data. (B) Percentage of correct answers for each question within each group. (C) Reaction times for each question within each group.

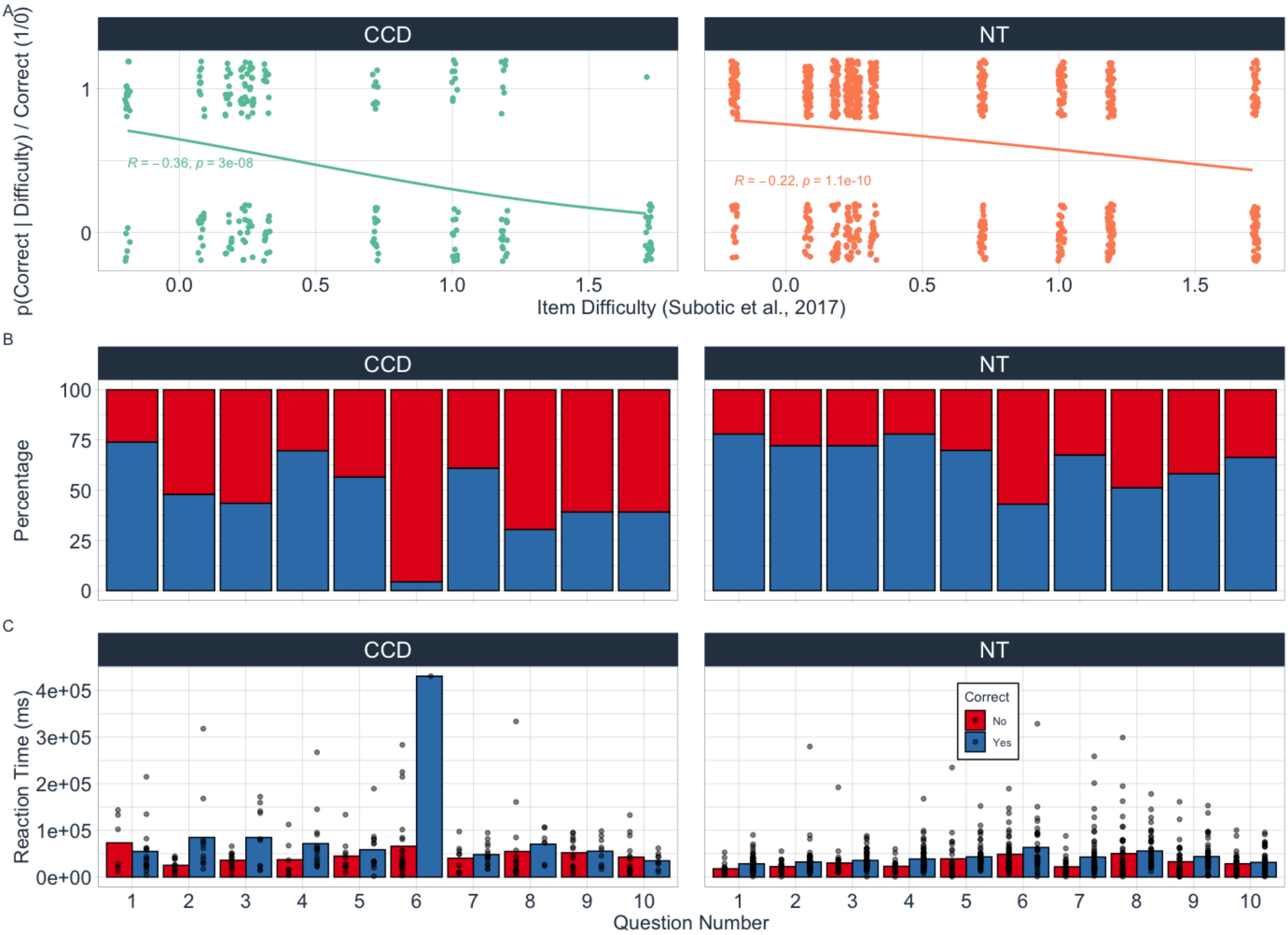

**Figure S3 Correlation and partial correlation matrices.**

X = non-significant association (alpha = 0.05).

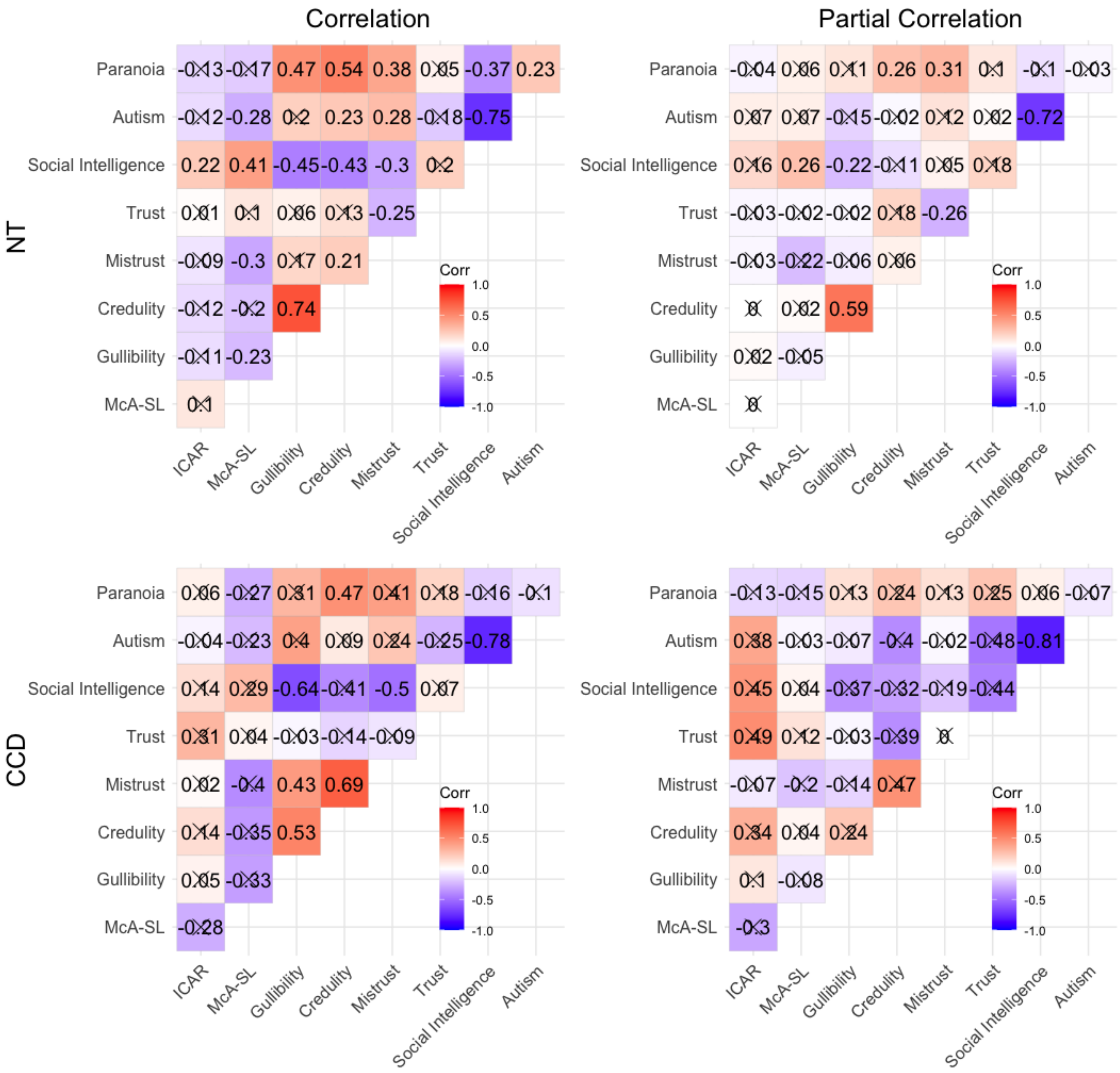

**Table S1 Descriptive statistics (mean [+/- sd]) for all measures completed by NT and CCD participants.**

Education = highest level of education achieved as of the current date. * = these figures only include participants in group two who completed the Social Intelligence and Autism Quotient scales in CCD (n = 20).

| **Variable** | NT (Sample who completed measures previously) | NT | CCD (Sample who completed measures previously) | CCD |
|---|---|---|---|---|
| ***n*** | 110 | 86 | 12 | 22 |
| **Age** | 29.13 [7.89] | 29.36 [7.60] | 51.75 [18.38] | 47 [17.26] |
| **Sex** | 38.2% Female | 40.2% Female | 50% Female | 59.10% Female |
| **Education (*n*)** | | | | |
| *Primary* | 0 | 1 | | 0 |
| *Secondary (e.g., High School Cert)* | 32 | 20 | | 12 |
| *Undergrad (e.g., BSc, MBBS)* | 55 | 42 | | 5 |
| *Postgrad (e.g., MSc, MA, PGc)* | 21 | 21 | | 5 |
| *Doctorate (e.g., PhD, DClinPsy)* | 2 | 2 | | 0 |
| **ICAR** | 6.66 [2.37] | 6.56 [2.31] | | 4.73 [2.62] |
| **Paranoia** | | | | |
| *Persecutory Ideation* | 13.2 [5.67] | 12.2 [4.94] | | 15.1 [8.46] |

| | | | | |
|---|---|---|---|---|
| *Social Reference* | 18.3 [7.15] | 16.0 [6.08] | | 15.7 [6.21] |
| **Gullibility** | | | | |
| *Insensitive* | 18.2 [6.49] | 17.9 [6.26] | 23.9 [7.95] | 22.8 [8.34] |
| *Persuadable* | 14.6 [6.73] | 13.1 [6.44] | 22.3 [8.51] | 20.2 [8.46] |
| **Epistemic Trust** | | | | |
| *Trust* | | 24.1 [4.85] | | 27.4 [4.71] |
| *Mistrust* | | 21.6 [4.29] | | 20.4 [5.39] |
| *Credulity* | | 12.9 [4.76] | | 18.6 [7.90] |
| **MacArthur Social Ladder** | | | | |
| *National* | 55.6 [20.16] | 52.5 [21.02] | | 51.1 [24.15] |
| *Community* | 48.1 [20.63] | 49.0 [19.71] | | 50.5 [28.64] |
| **Social Intelligence** | | 95.67 [23.46] | 79.78 [32.99] | 82.9 [21.08]* |
| **Autism** | | | | |
| *Short Scale* | | 65.5 [9.96] | 74.6 [14.10] | 75.6 [12.13]* |
| *Revised Scale* | | 34.9 [4.92] | 38.7 [7.54] | 41.0 [6.45]* |

### Table S2 Cronbach's alpha for each psychometric measure.

The original Cronbach alpha score for each paper is: ICAR (Condon & Revelle, 2014), Gullibility (Teunisse et al., 2020), Paranoia (Green et al., 2008; Freeman et al., 2020), Epistemic Trust (Campbell et al., 2021), Social Intelligence (Silvera et al., 2001; Grieve & Maher, 2013), Autism (Baron-Cohen et al., 2001; Hoekstra et al., 2011). MacArthur Social Ladder reliability analysis from previous papers is not available as the measure only comprises two items.

| Variable | Original Score | NT | CCD |
|---|---|---|---|
| **ICAR** | 0.68 | 0.65 | 0.63 |
| **Gullibility** | 0.92 | 0.91 | 0.87 |
| *Insensitive* | 0.87 | 0.84 | 0.75 |
| *Persuadable* | 0.88 | 0.88 | 0.83 |
| **Paranoia** | 0.90-0.95 | 0.91 | 0.94 |
| *Persecutory Ideation* | 0.90-0.97 | 0.87 | 0.93 |
| *Social Reference* | 0.90-0.95 | 0.85 | 0.85 |
| **Epistemic Trust** | 0.78 | 0.65 | 0.85 |
| *Trust* | 0.76 | 0.72 | 0.56 |
| *Mistrust* | 0.72 | 0.54 | 0.53 |
| *Credulity* | 0.81 | 0.72 | 0.87 |
| **MacArthur Social Ladder** | NA | 0.78 | 0.73 |
| **Social Intelligence** | 0.85-0.87 | 0.93 | 0.84 |
| **Autism** | 0.63-0.77 | 0.87 | 0.86 |
| *Short Measure* | 0.77-0.86 | 0.79 | 0.78 |
| *Revised Measure* | - | 0.57 | 0.69 |
